# SARS-CoV-2 virus culture from the upper respiratory tract: Correlation with viral load, subgenomic viral RNA and duration of illness

**DOI:** 10.1101/2020.07.08.20148783

**Authors:** Ranawaka APM Perera, Eugene Tso, Owen TY Tsang, Dominic NC Tsang, Kitty Fung, Yonna WY Leung, Alex WH Chin, Daniel KW Chu, Samuel MS Cheung, Leo LM Poon, Vivien WM Chuang, Malik Peiris

**Author notes:** **Address for correspondence:** Malik Peiris, School of Public Health, The University of Hong Kong, No 7 Sassoon Rd, Pokfulam, Hong Kong Special Administrative Region, China.

## Abstract

In 68 respiratory specimens from a cohort of 35 COVID-19 patients, 32 of them with mild disease, we found SARS coronavirus-2 virus culture and sub-genomic RNA was rarely detectable beyond 8 days after onset of illness although virus RNA by RT-PCR remained detectable for many weeks.

## Introduction

SARS-CoV-2 emerged in late 2019 to cause a pandemic of severe respiratory disease with major impact on global health and economies. Virus RNA may be detectable by RT-PCR long after clinical recovery, some patients remaining positive for three weeks or longer *(1,2)*. Similar findings were seen with SARS in 2003 *(3)*. A significant proportion of transmission occurs prior to and soon after onset of illness *(4)*. However, the duration of contagiousness after the onset of clinical symptoms remains poorly understood. This is relevant to determining policy on discharge of patients from containment in hospitals.

Viral RNA detection by RT-PCR does not prove the presence of infectious virus whereas culture isolation of virus is a better indication of contagiousness. Recent studies on experimentally infected hamsters showed efficient transmission of SARS-CoV-2 to contact hamsters on day 1 after challenge when virus culture was positive in the nasal washes but there was no transmission by day 6 when nasal washes were culture negative although viral load determined by RT-PCR was still high (>6.0 log10 RNA copies per mL) *(5)*. Thus virus culture may be a better surrogate for transmissibility. We attempted virus isolation in 68 specimens collected from 35 patients collected at different times after symptom onset to define the kinetics of culturable virus in upper respiratory specimens of patients with RT-PCR confirmed COVID-19 disease, those specimens with viral load ≥5 log_10_ also being examined for detection of sub-genomic viral RNA (sgRNA).

## Methods

Patients were diagnosed as COVID-19 by RT-PCR at individual hospitals. Follow up specimens were sent to the School of Public Health, The University of Hong Kong for virus culture and viral RNA load determination. Virus sgRNA was tested for in specimens with ≥5 log_10_ N gene copies per mL. Methods for virus N gene copy number quantification *(6)*, virus culture and sgRNA detection are detailed in the Appendix. The study was approved by the Research Ethics Committee of the Kowloon West Cluster reference No. KW/EX-20-039 (144-27) of the Hospital Authority of Hong Kong.

## Results

Sixty eight specimens from thirty five patients were studied. Patients with prolonged virus shedding (10 remaining virus RNA positive for >30 days) and those re-admitted because of RT-PCR positivity detected after discharge (N=6) were oversampled. Patient-age ranged from 17 to 75 years (median 38 years) and 44 of them were male (Table). Nine patients had underlying co-morbidities, including diabetes mellitus, hypertension, ischemic heart disease, atrial fibrillation, chronic obstructive airways disease, carcinoma of the lung, mild renal dysfunction and chronic hepatitis B carrier. None of the patients were immunocompromised. Three patients were asymptomatic, twenty nine patients had mild clinical illness (mild influenza-like illness symptoms, not requiring supplemental oxygen, or requiring < 3 liters / minute), two patients were critical (intubated, required ECMO or in shock) and one died. For asymptomatic patients, days after onset was estimated as days after first RT-PCR detection. Specimens submitted for virus culture were nasopharyngeal aspirates combined with throat swab (n=49), nasopharyngeal aspirate (n=2), nasopharyngeal swab combined with throat swab (n=3), nasopharyngeal swab (n=2), sputum (n=11) and saliva (n=1). The duration after onset of illness to specimen collection ranged from 1 to 67 days.

The median age of the culture positive and negative patients was 39 and 38 years, respectively. SARS-CoV-2 N gene copy number in the specimens ranged from 9.5 log_10_ copies per mL to undetectable (limit of detection: 10 copies per mL) (Figure 1), with the mean viral load in the culture positive and negative samples being 7.6 and 3.8 log_10_ copies per mL of specimen, respectively.

**Figure 1:**
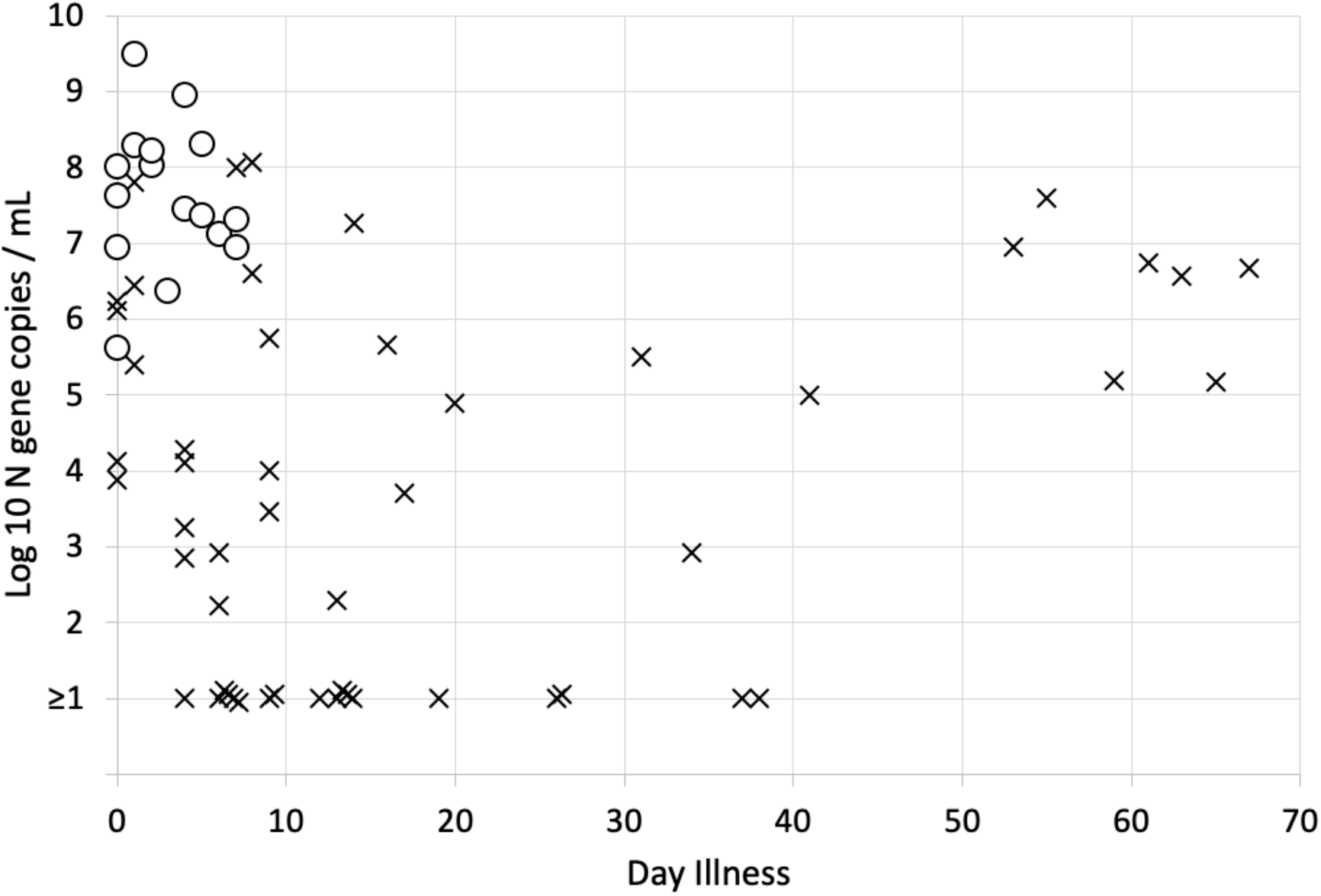
Viral RNA load, virus culture and days after onset of illness. Open circles indicate samples positive for virus culture and black crosses indicate virus culture negative samples in relation to days after onset of illness (X axis) and log 10 virus N gene copies / mL of specimen (Y axis). The limit of detection of the viral N gene RNA was 1 log10 copies / mL

Virus was cultured from 12 of 17 specimens with virus load ranging from 7.0-9.5 log_10_ copies per mL of specimen, 3 of 11 specimens with viral load ranging from 6.0-6.99 log_10_ copies per mL, 1 of seven specimens with viral load ranging from 5.0-5.99 log_10_ copies per mL and from none of 33 specimens with viral load <5log_10_ copies per mL (Table) (Figure 1). Virus isolation was successful in 8 of 15 specimens collected in the first two days after onset of symptoms, 3 of 8 specimens on day 3-4 after onset, 3 of 10 specimens on day 5-6 after onset, 2 of 5 on days 7-8 after onset and 0 (0%) of 30 specimens collected from day 9 or later after onset of illness (Table) (Figure 1).

Sub-genomic RNA (sgRNA) provides evidence of replicative intermediates of the virus rather than residual viral RNA. Detection of virus sgRNA was attempted in 33 of the 35 the specimens with viral load ≥5.0 log10 virus genome copies per mL of clinical specimen, two specimens had insufficient specimen for this additional testing. Of 33 specimens tested for both sgRNA and virus culture, both were positive in 12 (36.3%) specimens, both were negative in 36.4%, sgRNA was positive and culture negative in 7 (21.2%) while culture was positive and sgRNA was negative in 2 (6.1%), the association between virus culture and sgRNA being significant (Chi squared test with Yates correction p=0.014). Virus sgRNA was detectable in 18 (81.8%) of 22 specimens collected 8 days or less after symptom onset and in 1 (9.1%) of 11 specimens collected 9 or more days after onset of disease (Chi squared test with Yates correction p=0.0003) (Figure 2).

**Figure 2:**
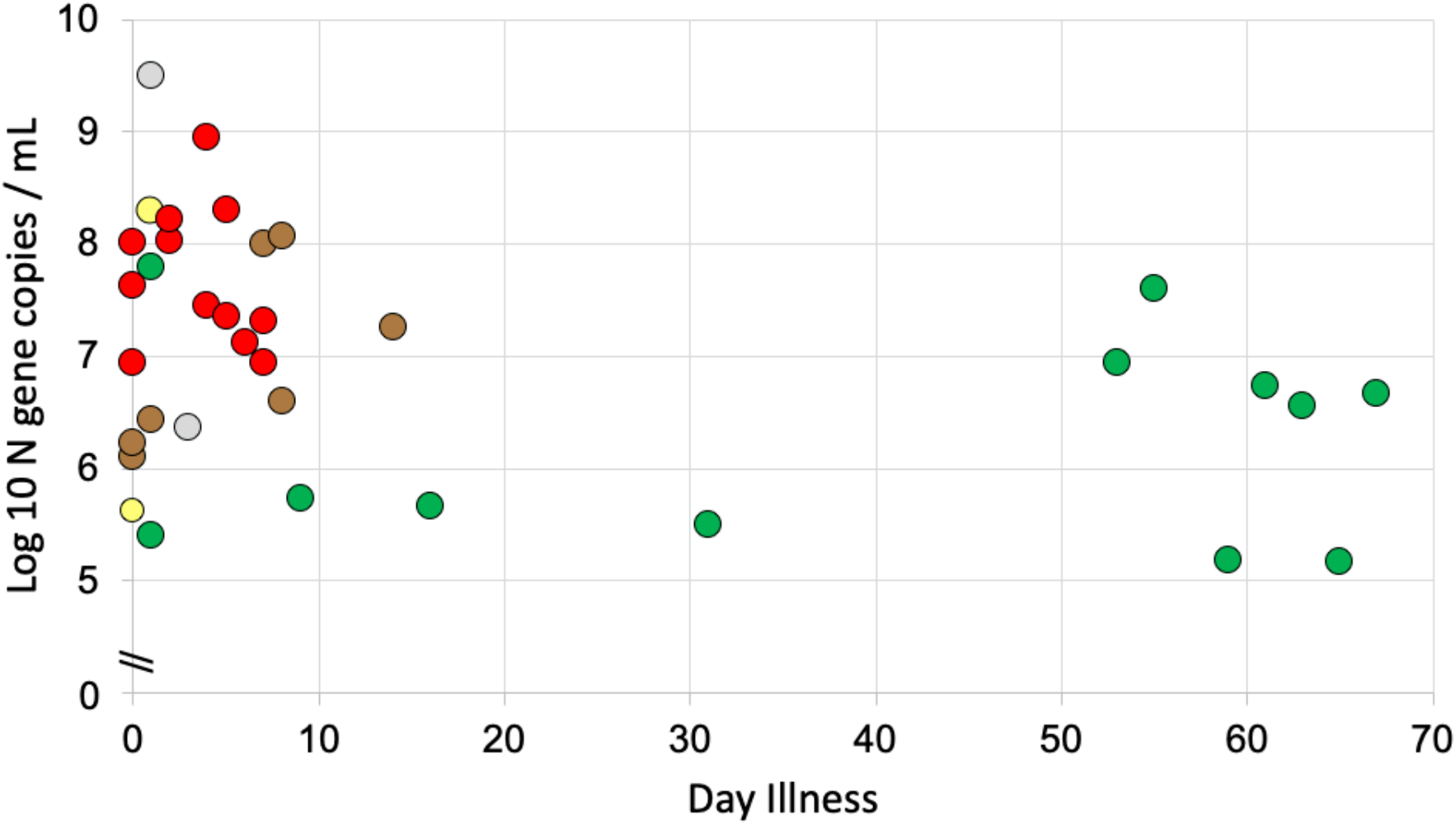
Viral RNA load, virus culture, subgenomic virus RNA (sgRNA) in relation to days after onset of illness. Red dots indicate samples positive for both culture and sgRNA detection. Green dots were specimens negative for both virus culture and sgRNA, yellow was virus culture positive and sgRNA negative while brown indicated virus culture negative and sgRNA positive. Grey dots indicate samples with virus culture positive but without sgRNA data because of insufficient specimen.

**Table 1.** Comparison of patients and clinical specimens that were positive or negative for virus isolation.

| Patient characteristics | Culture positive<br>N=16 | Culture negative<br>N=52 | Total<br>N=68 | Statistical<br>significance |
| --- | --- | --- | --- | --- |
| Patient (n=35) characteristics |  |  |  |  |
| Asymptomatic | 1 | 2 | 3 |  |
| Median age years<br>(range) | 39 (21-73) | 38 (17-75) | 38 |  |
| Co-morbidities | 5 | 4 | 9 |  |
| Clinical specimen (n=68) characteristics |  |  |  |  |
| Mean Log<sub>10</sub> viral<br>load/mL | 7.6 | 3.8 |  | Mann<br>Whitney<br>p<0.001 |
| Viral load Log<sub>10</sub> ranging from |  |  |  |  |
| 7.0 to 9.5 | 12 (75%) | 5 (10%) | 17 (25%) | Chi squared<br>p< 0.00001 |
| 6.0 to 6.99 | 3 (19%) | 8 (15%) | 11 (16%) |  |
| 5.0 to 5.99 | 1 (6%) | 6 (12%) | 7 (10%) |  |
| <5.0 | 0 (0%) | 33 (63%) | 33 (49%) |  |
| Days after onset of illness at which sample was collected |  |  |  |  |
| 1 to 2 | 8 (53%) | 7 (13%) | 15 (22%) | Chi squared<br>p< 0.001 |
| 3 to 8 | 8 (35%) | 15 (29%) | 23 (34%) |  |
| 9 to 67 | 0 (0%) | 30* (58%) | 30 (44%) |  |

\* 6 of these were readmissions because of detection of virus RNA after discharge from isolation.

## Discussion and Conclusions

In a cohort of predominantly mild COVID-19 patients, our findings suggested that virus isolation and sgRNA detection was possible mainly within the first eight days after onset of illness and from specimens with ≥6 log10 virus N gene copies per mL of clinical specimen. Although there were five specimens with viral load ≥6 log10 virus N gene copies per mL later than 50 days after onset of illness, none of these yielded culturable virus or virus sgRNA. Thus viral load itself was not predictive of presence of infectious virus or sgRNA. We did not carry out serological testing in parallel with viral culture but data on a larger cohort of patients followed up in Hong Kong showed that most patients had detectable (titer ≥1:10) plaque reduction neutralization antibody titres after day 9 of illness *(7)*. There are two other studies of virus culture on mild or moderately ill patients and virus culture was successful only within the first 9 days after onset of illness *(8,9)*. Patients who are severely ill and immunocompromised may shed infectious virus for much longer periods of time (10).

Taken together previous data, our findings suggest that patients with mild or moderate illness may be less contagious after eight days after symptom onset. Mildly ill patients who have clinically recovered and not immunocompromised may be discharged from containment after 9 days or more after symptom onset, while ensuring that they are not being discharged into settings with other highly vulnerable individuals (e.g. old age care homes). These findings support the recent change in the WHO guidelines for releasing COVID-19 patients from isolation; viz. 10 days after symptom onset and at least 3 additional days without symptoms *(11)*.

## Data Availability

The authors confirm that the data supporting the findings of this study are available within the article.

## Acknowledgments

This work was supported by the US National Institutes of Health (contract no. HHSN272201400006C) and a Natural Science Foundation of China (NSFC)/Research Grants Council (RGC) Joint Research Scheme (N_HKU737/18).

## Author Bio

Ranawaka APM Perera is a research assistant professor at the School of Public Health, The University of Hong Kong with a research interest in emerging virus infections including influenza, MERS coronavirus and SARS coronavirus-2.

